# Sub-optimal Neutralisation of Omicron (B.1.1.529) Variant by Antibodies induced by Vaccine alone or SARS-CoV-2 Infection plus Vaccine (Hybrid Immunity) post 6-months

**DOI:** 10.1101/2022.01.04.22268747

**Authors:** Guruprasad Medigeshi, Gaurav Batra, Deepika Rathna Murugesan, Ramachandran Thiruvengadam, Souvick Chattopadhyay, Bhabatosh Das, Mudita Gosain, Ayushi, Janmejay Singh, Ananthraj Anbalagan, Heena Shaman, Kamal Pargai, Farha Mehdi, Soon Jyoti Das, Namrata Kahlon, Savita Singh, Pallavi Kshetrapal, Nitya Wadhwa, Anil K Pandey, Shinjini Bhatnagar, Pramod Kumar Garg

**Affiliations:** Translational Health Science and Technology Institute, Faridabad, Haryana, India; ESIC Medical College and Hospital, Faridabad, Haryana, India

**Author notes:** Corresponding author Prof. Pramod K Garg MD, Translational Health Science and Technology Institute, Faridabad 120001, Haryana, India.

## Abstract

**Background:** Rapid expansion of the omicron SARS-CoV-2 variant of concern despite extensive vaccine coverage might be related to decreased neutralising ability of vaccine induced antibodies. The neutralising ability of different vaccines with or without natural SARS-CoV-2 infection against omicron is however not well known.

**Methods:** We tested the ability of vaccine and natural infection induced antibodies to neutralise omicron variant in a live virus neutralisation assay. Four groups of individuals were included: (i) complete vaccination with ChAdOx1 nCoV-19 (n=20), (ii) complete vaccination with ChAdOx1 nCoV-19 plus prior SARS-CoV-2 infection during the delta variant driven surge (n=20), (iii) complete vaccination with inactivated whole virus vaccine (BBV152) (n=20), (iv) complete vaccination with BBV152 plus prior SARS-CoV-2 infection (n=20). Primary outcome was fold-change in the virus neutralisation ability of plasma against the omicron variant compared with ancestral and delta variant.

**Findings:** The neutralisation geometric mean titre (GMT) was 384 (95% CI: 662, 223) against the ancestral virus with BBV152 vaccination alone and 383 (95% CI: 709, 207) with ChAdOx1 nCov-19 vaccination alone. The corresponding values for hybrid immunity groups were 795 (95% CI: 1302, 486) and 1424 (95% CI: 2581,786) respectively. Against the omicron variant, only 5 out of 20 in both BBV152 and ChAdOx1 nCoV-19 vaccine only groups, 5 out of 19 in BBV152 plus SARS-CoV-2 infection group, and 9 out of 20 in ChAdOx1 nCoV-19 plus SARS-CoV-2 infection group exhibited neutralisation titres above the lower limit of quantification (1:20) suggesting better neutralization in those with prior infection. The 50% neutralisation against ancestral strain and omicron demonstrated strong correlation with anti-RBD IgG levels [Pearson r: 0.94 (0.91, 0.96) p: <0.001 and 0.92 (0.88, 0.95) p:<0.001 respectively].

**Interpretation:** Omicron variant shows significant reduction in neutralising ability of both vaccine induced and hybrid immunity induced antibodies which might explain immune escape and high transmission even in the presence of widespread vaccine coverage.

**Funding:** DBT, India; GIISER-BMGF, USA

**Research in context:** *Evidence before this study:* The Omicron variant of SARS-CoV-2 is fast becoming the dominant circulating strain world-wide. We did a literature search on PubMed between 01 November 2020 to 04 January 2022 using the terms “Omicron” and “neutralisation” and found 11 results for virus neutralisation against omicron by vaccine/natural infection induced antibodies. We identified two published and one preprint articles relevant to omicron virus neutralisation using live virus neutralization. Preliminary reports suggest that omicron variant is significantly less susceptible to *in-vitro* neutralisation by antibodies among recipients of mRNA vaccines (BNT162b2 and mRNA-1273), adenovirus vectored vaccine (ChAdOx1 nCoV-19 vaccines) and no virus neutralization was observed in subjects who received Coronavac (inactivated virus vaccine). Data regarding immune escape among those with natural SARS-CoV2 infection and vaccination are not available.

*Added value of this study:* We report here that the proportion of neutralisers (those who demonstrated a FRNT50 titre >1:20) was significantly reduced against the omicron variant as compared to the ancestral and delta variant. The geometric mean titre of neutralisation among the vaccinated individuals without a history of previous natural infection was significantly reduced against the omicron variant as compared with ancestral and delta variants. The titres among the those with a history of previous infection also followed the same pattern, but the neutralising ability was better in them than those who did not have previous infection.

*Implications of all the available evidence:* Omicron variant of SARS-CoV-2 is capable of escaping immunity provided by currently available vaccines and even natural infection due to significant mutations in its spike protein. The drop in neutralisation might be alarming, but the real-world impact of these reduced neutralisation titres on major public health indices like hospitalisation rates and mortality rates have to be interpreted along with the other factors such as inherent pathogenicity of the variant, immunization uptakes and seroprevalence from natural infection in different geographical regions and the expected role of cellular immune responses to the variant. Our data may guide policy on booster vaccination to deal with an impending public health emergency as a result of surge in omicron cases.

---

Severe acute respiratory syndrome coronavirus 2 (SARS-CoV-2) has affected >280 million people and claimed >5.4 million lives worldwide.(1) The most effective strategy to contain the pandemic is vaccination. However, the emergence of variants of concern (VoC) due to mutations in the virus has led to reduced vaccine effectiveness. After the massive surge due to the delta (B·1·617·2) VoC, the emergence and rapid spread of the Omicron (B.1.1.529) VoC has further caused panic around the world. The virus enters the human respiratory epithelial cells by binding the receptor-binding domain (RBD) of its spike (S) protein with the human angiotensin converting enzyme-2 (ACE2) receptors. The predominant mechanism of protection afforded by the vaccines is through generation of neutralising antibodies against the RBD that help block viral entry into host cells. The VoC harbouring mutations in the RBD may lead to a decreased neutralising ability of the vaccine-derived antibodies as has been shown against VoCs such as the alpha, beta, gamma and delta variants.(2,3) Omicron has 30 amino acid substitutions, 3 deletions, and 1 insertion in its spike protein (15 substitutions in RBD alone) and thus may escape vaccine induced immunity.(4) Preliminary data derived from two recent studies from South Africa and the United Kingdom have shown markedly reduced ability of the serum from vaccinated people to neutralize Omicron in live virus neutralisation assays. These data pertain to a small sample size of people vaccinated with either mRNA vaccine (BNT162b2; Pfizer-BioNTech) or adenovirus vectored (Oxford-AstraZeneca) vaccines.(5,6) The ability of the sera to neutralise Omicron from people vaccinated with inactivated virus vaccine or those with hybrid immunity following natural SARS-CoV-2 infection plus vaccination is still unclear. This assumes significance because of the high prevalence of asymptomatic infection in many countries as detected by serosurveys.(7,8) A hybrid immunity is likely to provide better protection and thus help guide policy about vaccinating or boosting naturally infected people. India’s vaccination program is driven by ChAdOx1 nCoV-19 vaccine (Covishield, Serum Institute of India, Pune, India) and an inactivated whole virus vaccine (BBV152; Covaxin, Bharat Biotech Immunologicals Limited, Hyderabad, India). India witnessed a massive surge due to the delta (B·1·617·2) VoC during April-May 2021.(9) Serosurvey following this surge showed a high prevalence (69.2 %) of IgG antibodies suggesting widespread infection albeit mostly asymptomatic.(7) In the present study, we tested the ability of antibodies to neutralise the omicron variant among people with vaccination alone or vaccination plus natural infection induced immunity.

## Methods

### Study Participants

The participants were derived from an ongoing Department of Biotechnology (DBT) consortium for COVID-19 Research cohort developed by Translational Health Science and Technology Institute, Faridabad, India in collaboration with hospitals in Delhi National Capital region, particularly, Employee State Insurance Corporation Medical College and Hospital, Faridabad. We randomly sampled ChAdOx1 nCoV-19 and BBV152 vaccine recipients from the cohort and categorised them in the following four groups: (i) vaccination with two doses of ChAdOx1 nCoV-19 alone, (ii) vaccination with two doses of ChAdOx1 nCoV-19 along with an RT-PCR confirmed natural infection during the delta variant driven surge, (iii) vaccination with two doses of BBV152 alone, (iv) vaccination with two doses of BBV152 along with an RT-PCR confirmed natural infection during the delta variant driven surge. Written informed consent was obtained before enrolling the participants.

### Data collection

Specific details were obtained from the DBT Consortium cohort database and during interviews by trained research staff on the vaccination status of the participants including the name of the vaccine, number of doses, date and place of administration, and clinical manifestations of COVID-19 among those with natural infection in the past. We defined complete vaccination when the participant had completed at least 14 days after the second dose of the vaccine. The dates of immunization were documented from the certificate of immunization issued by the Government of India. Their blood samples were collected and processed for plasma and peripheral blood mononuclear cells.

### Ethical Approval

The studies were approved by the Institute Ethics Committees of the partnering institutions.

#### Anti-SARS-CoV-2 IgG antibody detection

Anti-RBD IgG concentrations in binding antibody units/ml (BAU/ml) in the serum samples were determined by enzyme-linked immunosorbent assay (ELISA) using internal control calibrated against the first WHO international standard for anti-SARS-CoV-2 immunoglobulin (code 20/136).(10) In addition, anti-nucleocapsid antibodies were detected using a qualitative ELISA. The detailed methods are provided in the supplementary material.

#### Live Virus Focus Reduction Neutralisation Assay

Virus neutralisation assay was performed as described previously with minor changes in the incubation periods.(11) Briefly, plasma samples were serially diluted from 1:20 to 1:640 and virus neutralization was tested in Vero E6 cells. Cells were incubated for 24 hours for ancestral (B.1) and Delta (B.1.617.2) variants and for 32 hours for Omicron variant.(12) All virus stocks used in this study were propagated in Calu-3 cells (American Type Culture Collection). Total RNA sample of the culture was processed for whole genome sequencing using an Illumina MiSeq sequencing platform to confirm the variant. Sequencing reads (>Q30) were assembled by rnaSPAdes pipeline. Phylogenetic Assignment of Named Global Outbreak Lineages (PANGOLIN) program version 3.1.11 with pangoLEARN 2021-09-17 were used for lineage prediction. Microplaques were quantified by AID iSPOT reader (AID GmbH, Strassberg, Germany). 50% neutralisation values were calculated with four-parameter logistic regression using GraphPad Prism (version 9.3.1) software. All virus-related experiments were performed in a biosafety level-3 lab.

### Outcome measure

Our primary outcome was fold-change in the virus neutralisation ability of the plasma against the omicron variant as compared with ancestral virus and delta variant. A secondary outcome was correlation of the serum IgG titres with the neutralization geometric mean titre.

### Statistical analysis

We report the proportion of individuals whose plasma demonstrated in vitro live virus neutralization (defined as FRNT50 ≥20) against ancestral, delta and omicron variants, and compared the geometric mean titre (GMT) for each variant with the GMT of ancestral virus considered as the reference, using mixed effects analysis and Dunnett’s multiple comparison test. For this analysis, those who demonstrated FRNT50 <20 were considered to have a titre of 10. A neutralization GMT of 1:20 or more was taken as protective.(13) The FRNT50 of the participants was correlated with quantitative anti-RBD IgG antibody levels (BAU/ml) using Pearson’s correlation coefficient. The ELISA values between the vaccine only and vaccine with infection group, and anti-nucleocapsid antibody positive and negative groups were tested using Mann Whitney U test, with p-value <0.05 considered as significant. To evaluate whether type of vaccine and past SARS-CoV-2 infection were independent predictors of neutralisation titres against the omicron variant, we constructed a multivariable regression model with FRNT50 titre as the dependent variable and age, sex, type of vaccine and a history of RT-PCR positive SARS-CoV-2 infection as independent variables.

## Results

### Participants

We included a total of 80 participants – 20 each with ChAdOx1 nCoV-19 and BBV152 vaccination alone, and 20 each with ChAdOx1 nCoV-19 vaccine plus natural infection and BBV152 vaccine plus natural infection. Their median age was 58 years (IQR: 48, 65); 32% were females. The median duration from the second dose of vaccine was 234 days (IQR: 210, 274) and the median duration after the natural infection was 224 days (IQR: 199, 238). The clinical characteristics of the study participants are provided in Table 1.

**Table 1.**
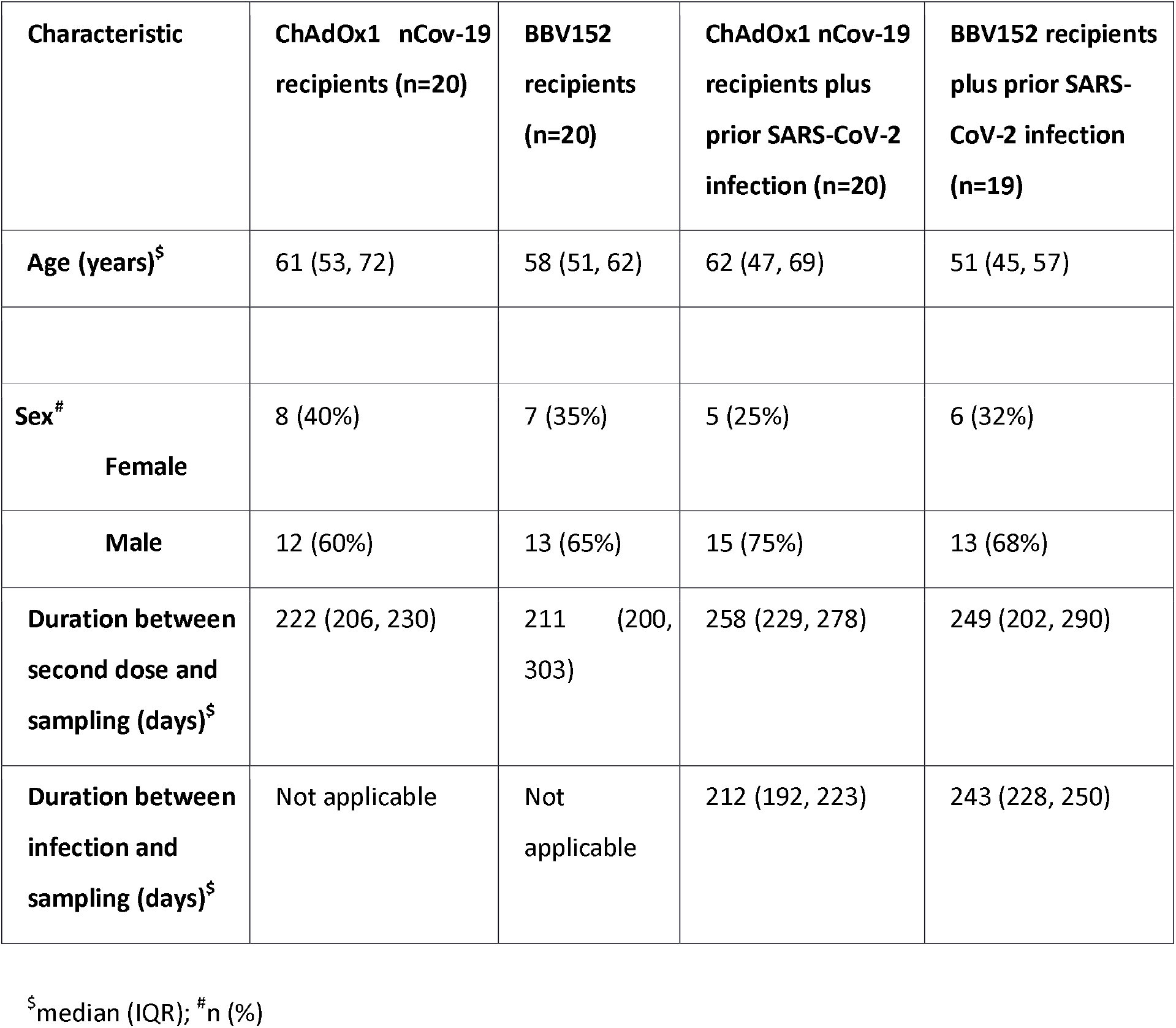
Clinical characteristics of the participants.

#### Serum anti-RBD IgG Antibody Titres

The median serum anti-RBD IgG antibody titre was 243.5 (IQR 111.3, 544.4) BAU/ml (one of the samples with a very high value of 85443.9 BAU/ml was considered an outlier and removed from analysis). The prevalence of anti-nucleocapsid antibodies was 1 of 20 ChAdOx1 nCoV-19 vaccination alone participants, 11 of 20 ChAdOx1 nCoV-19 vaccination plus infection participants, 8 of 20 BBV152 vaccination alone participants, and 18 of 20 BBV152 vaccination plus infection participants (Table S1). The median anti-RBD IgG titres were higher in the vaccination plus natural infection [403.9 (IQR 164.6, 794.8) BAU/ml] and anti-nucleocapsid IgG positive group [380.7 (IQR 178.0, 729.9)] BAU/ml compared to the vaccine alone group [150.6 (IQR 81.9, 298.5) BAU/ml] (p-value <0.001) and anti-nucleocapsid IgG negative group [161.6 (IQR 82.8, 338.4) BAU/ml] (p-value 0.003){Table S2}.

#### Live Virus neutralisation ability of plasma

The neutralisation geometric mean titre (GMT) was 384 (95% CI: 662, 223) against the ancestral virus with BBV152 vaccination alone. Similar GMT values, 383 (95% CI: 709, 207), were obtained with ChAdOx1 nCov-19 vaccination alone (Figure 1, Table 2). The corresponding values for hybrid immunity groups were 795 (95% CI: 1302, 486) and 1424 (95% CI: 2581,786) respectively. The GMT showed a 2.2 to 3.4-fold reduction against the delta variant (B.1.617·2) as compared with the ancestral virus in all the four groups (Table 2). Against the omicron variant, only 5 out of 20 in both the BBV152 and ChAdOx1 nCoV-19 vaccine only groups, 5 out of 19 in BBV152 plus SARS-CoV-2 infection group, and 9 out of 20 in ChAdOx1 nCoV-19 plus SARS-CoV-2 infection group exhibited neutralisation titres above the lower limit of protection (1:20) (Figure 1, Table 2). We observed a 25 to 27-fold reduction in GMT values against the Omicron variant in the vaccination alone groups and 52 to 54-fold reduction in the hybrid immunity groups compared with the ancestral virus. As expected, subjects who were infected with SARS-CoV-2 post-vaccination showed an increase in titres against the delta variant in both BBV152 and ChAdOx1 nCoV-19 vaccinated groups. The GMT for delta variant in these cases increased from 174 (95% CI: 291, 104) to 318 (95% CI: 520, 194) in the BBV152 vaccination and from 111 (95% CI: 195, 64) to 451 (95% CI: 760, 268) in the ChAdOx1 nCoV-19 vaccination group. Interestingly, natural SARS-CoV-2 infection, most likely due to the delta variant, led to an increase in titres against the ancestral virus, from GMT 384 (95% CI: 662, 223) to 795 (95% CI: 1302, 486) in the BBV152 group and from GMT 383 (95% CI: 709, 207) to 1424 (95% CI: 2581,786) in the ChAdOx1 nCoV-19 group (Figure 1, Table 2). However, the GMT declined significantly in the case of omicron. The FRNT50 titre against the ancestral and the omicron variant demonstrated strong correlation with anti-RBD IgG levels [Pearson r: 0.94 (0.91, 0.96) p: <0.001 and 0.92 (0.88, 0.95) p:<0.001 respectively] (Figure 2). Past SARS-CoV-2 infection during the delta surge was independently associated with 26.06 (95%CI: 2.15, 69.97) neutralization increase against the omicron as compared to those uninfected after adjusting for age, sex and type of vaccine. The type of vaccine had no influence on the neutralisation titre against the Omicron (Table 3).

**Figure 1:**
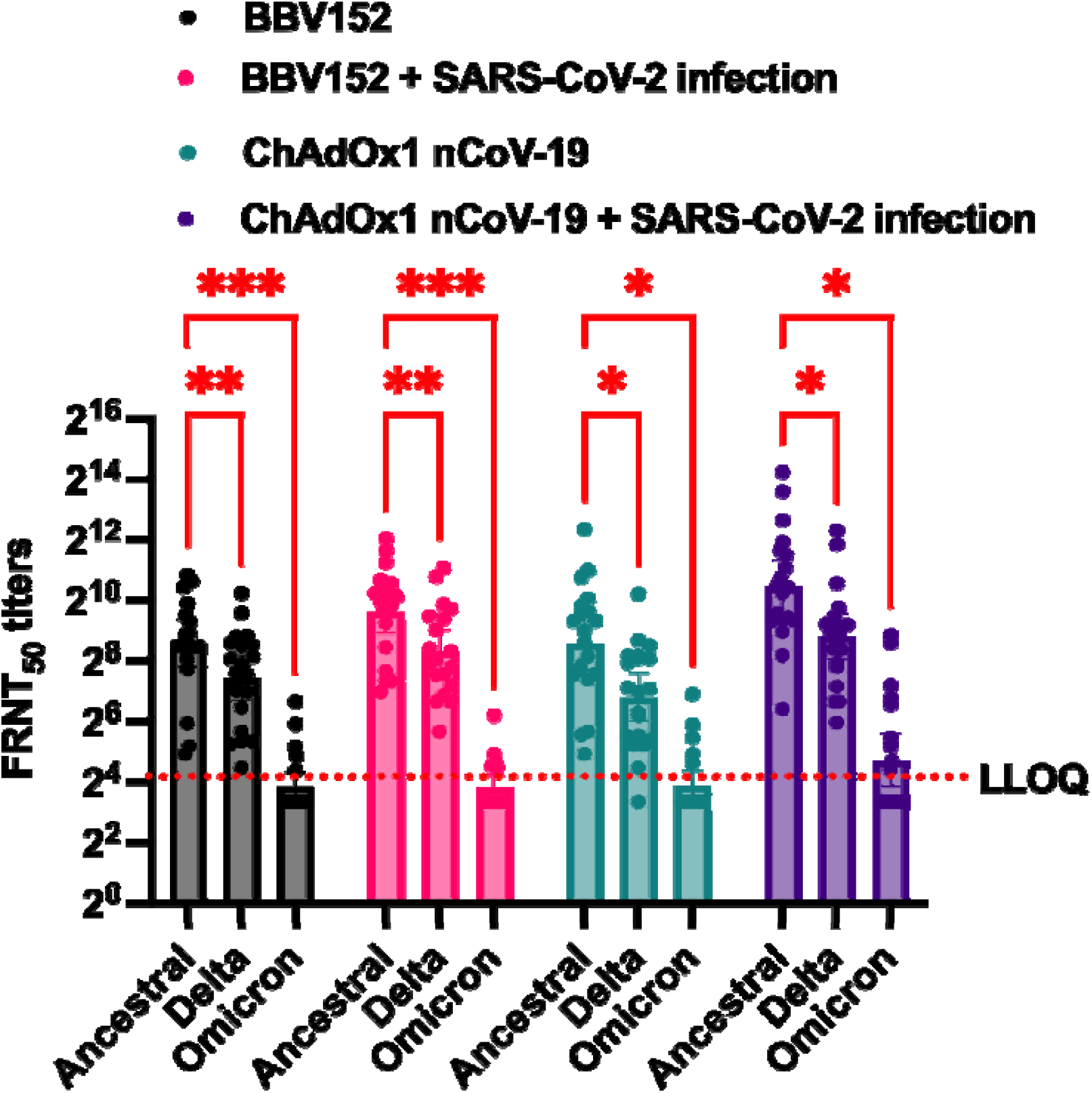
Virus neutralization assay (N=79) The dotted line represents FRNT50 of 20, which was considered as lower limit of quantification. Statistical test – Analysis of variance was performed with mixed effects method with ancestral virus neutralisation titres considered as the reference adjusted using Dunnett’s multiple comparison test. LLOQ = lower limit of quantification (1:20)

**Table 2.** Neutralization geometric mean titres of participants against SARS-CoV-2 variants.

|  | GMT (95% CI) by NT<sub>50</sub> |  |  |  |  |  |  |
| --- | --- | --- | --- | --- | --- | --- | --- |
| Type of participant | N<sup>1</sup> | Virus type |  |  | Fold Reduction Anc/Delta | Fold Reduction Anc/OMC | Fold Reduction Delta/OMC |
|  |  | Ancestral | Delta | Omicron |  |  |  |
| BBV152 | 20 | 384 (662, 223) | 174 (291, 104) | 14 (20,10) | 2.2 | 27.4 | 12.4 |
| BBV152 plus SARS-CoV-2 infection* | 19 | 795 (1302, 486) | 318 (520, 194) | 14 (19, 11) | 2.5 | 56.8 | 22.7 |
| ChAdOx1 nCov-19 | 20 | 383 (709, 207) | 111 (195, 64) | 15 (21, 10) | 3.5 | 25.5 | 7.4 |
| ChAdOx1 nCov-19 plus SARS-CoV-2 Infection* | 20 | 1424 (2581, 786) | 451 (760, 268) | 26 (49, 14) | 3.2 | 54.8 | 17.3 |

Anc: ancestral
<sup>1</sup>Number of participants whose plasma 50% virus neutralization titres (NT50) were estimated by focus reduction neutralization test against specific variants
Mean value of ancestral-type was considered as Control for comparison.
GMT – Geometric mean titre; NT50 – 50% neutralization in Focus Reduction Neutralization Test
\* SARS-CoV2 infection during delta (B.1.617.2) driven surge

**Figure 2.**
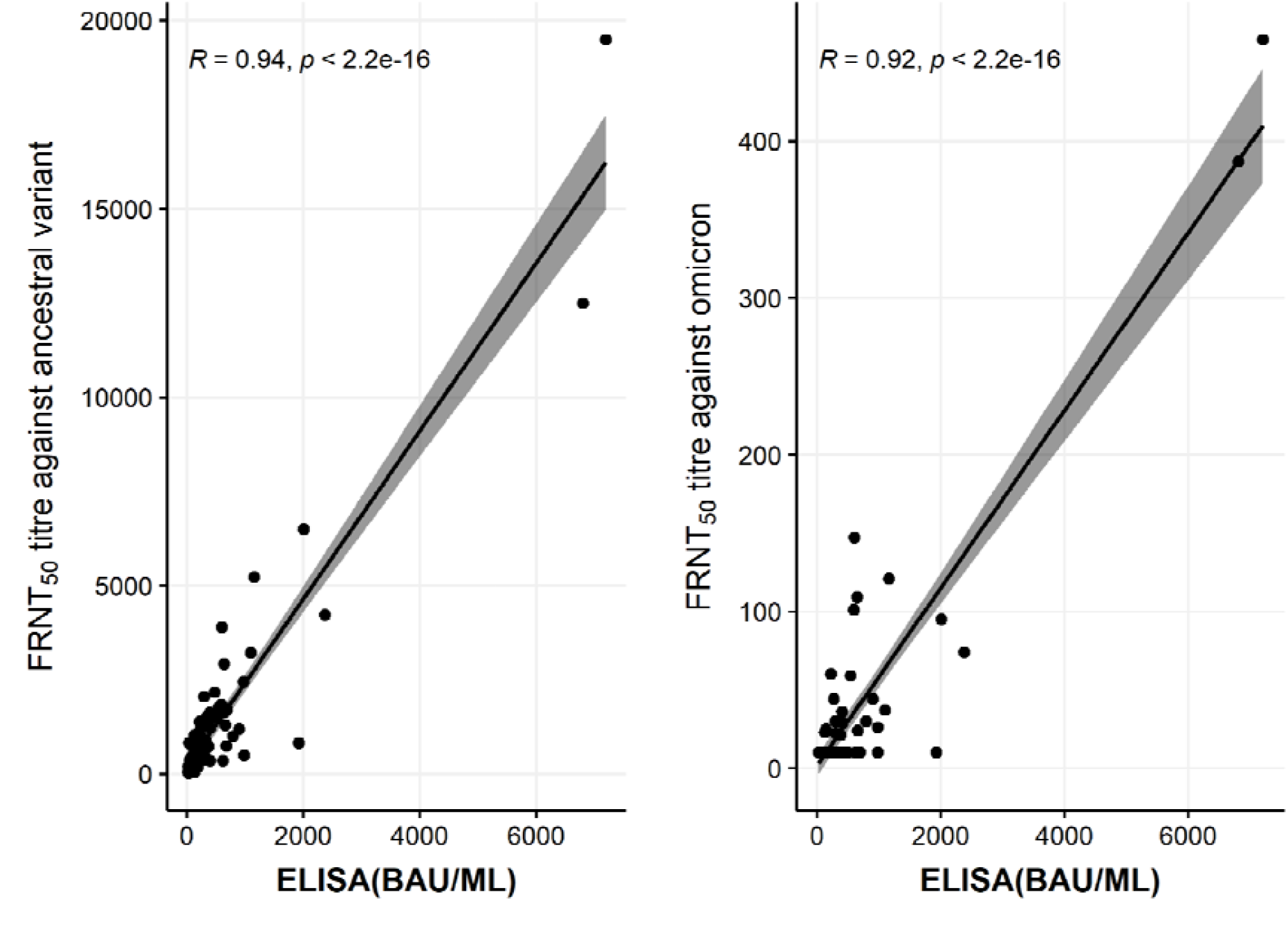
Correlation between FRNT50 titre against ancestral virus and omicron variant, and anti-RBD IgG ELISA levels (N=79)

**Table 3.**
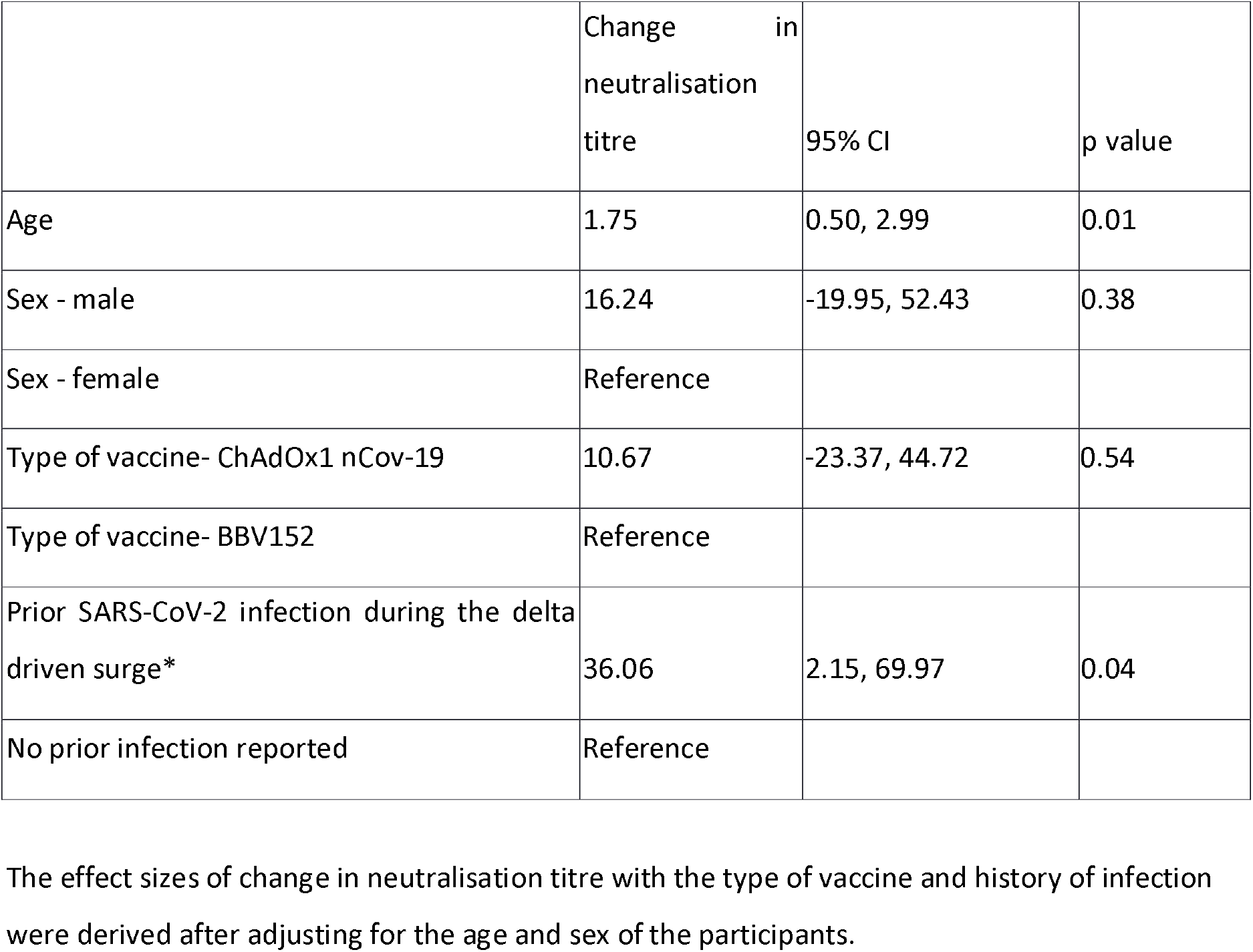
Association between type of vaccine and prior SARS-CoV-2 infection and neutralising titres against Omicron variant (N=79)

|  | Change in<br>neutralisation<br>titre | 95% CI | p value |
| --- | --- | --- | --- |
| Age | 1.75 | 0.50, 2.99 | 0.01 |
| Sex - male | 16.24 | -19.95, 52.43 | 0.38 |
| Sex - female | Reference |  |  |
| Type of vaccine- ChAdOx1 nCov-19 | 10.67 | -23.37, 44.72 | 0.54 |
| Type of vaccine- BBV152 | Reference |  |  |
| Prior SARS-CoV-2 infection during the delta driven surge* | 36.06 | 2.15, 69.97 | 0.04 |
| No prior infection reported | Reference |  |  |

The effect sizes of change in neutralisation titre with the type of vaccine and history of infection were derived after adjusting for the age and sex of the participants.

## Discussion

In the present study, recipients of BBV152 and ChAdOx1 nCoV-19 vaccines demonstrated significantly reduced in vitro virus neutralisation against the omicron variant regardless of the past infection with SARS-CoV-2. Mutations in the RBD region of the spike protein may cause compromised binding with the neutralising IgG antibodies and thus the reduced neutralisation observed in the earlier studies as well.(5,6,14–18) There was no difference in the neutralisation between the two types of vaccines tested in the present study. However, the neutralisation titres for the omicron were better among those vaccine recipients (particularly ChAdOx1 nCoV-19 vaccine), who had a natural infection during the delta variant led surge. Past infection was capable of boosting the antibody titres against the ancestral virus. A recent report has shown that individuals infected with the omicron variant demonstrated an enhanced antibody response against the delta variant suggesting that there could still be shared antibody epitopes between the omicron and other variants mainly pan-Sarbecovirus neutralizing antibodies binding outside the receptor binding motif region.(19,20)

It appears that anti-nucleocapsid antibodies induced by the whole inactivated virus BBV152 vaccine wane over 6 months as we could only detect these in 8 out of 20 participants in the BBV152 vaccination alone group. This is consistent with an earlier report of waning anti-nucleocapsid antibodies after natural infection.(21) Interestingly, 18 of 20 BBV152 vaccination plus infection group had anti-nucleocapsid antibodies compared to 11 of 20 ChAdOx1 nCoV-19 vaccination plus infection group suggesting boosting of anti-nucleocapsid IgG response after infection in the BBV152 vaccinated individuals. The dynamics of anti-nucleocapsid antibodies are important for population serosurvey to detect past infection.

The partial loss of neutralizing ability against Omicron has been demonstrated previously. Of particular interest to our population is the data from the UK that showed that just 1 out of 22 participants, whose samples were collected after 28 days from their second dose of ChAdOx1 nCoV-19 vaccine, demonstrated neutralisation against the omicron. Our participants who were sampled after six months from their second dose demonstrated a marginally better neutralisation potential against the omicron with 5 out of 20 participants in each vaccine group showing neutralisation, possibly due to prior but undetected SARS-CoV-2 infection. The UK data showed a 29-fold reduction against the omicron in the BNT162b vaccinated sera. In the present study, the fold reduction against the omicron variant was of the order of 25 to 56. Comparative fold reduction in the neutralising ability of vaccinated and hybrid immunity plasma was largely due to a significant increase in the serum IgG antibodies titres which are primarily generated against the ancestral virus. Since the omicron variant has a much different RBD with 30 substitutions, 1 insertion and 3 deletions in its spike protein, it is expected that the neutralising ability of the antibodies will be significantly compromised. However, from the point of view of vaccine/natural infection induced immunity against infection/re-infection, it is important to understand that a minimum virus neutralising ability is required for optimal protection. It has been shown that a 1:20 neutralisation titre at which 50% virus neutralisation can be achieved is expected to provide 80% vaccine effectiveness against the infection.(13) Therefore, we believe that it is more important to report what percentage of vaccinated/hybrid immunity people are able to neutralise the omicron variant at that GMT. Fold change showing very high reduction compared to the ancestral virus might be misleading. Our data should be interpreted keeping in mind that the mean age of the participants was 58 years with 47% of participants >60 years of age and the time elapsed after complete vaccination was around 7 months and after natural infection was also around 7 months.

Marked reduction in the neutralising ability of vaccines might translate into much reduced vaccine effectiveness against the omicron. However, while neutralising antibodies are important to prevent virus entry into the host cells, an important defence mechanism is the cellular immune responses against the virus. It has been shown that CD4+ and CD8+ T-cell immune responses are important for controlling SARS-CoV-2 infection.(22) This seems to be true for the omicron variant too as demonstrated by comparable and maintained CD4 and CD8 T cell response against the spike proteins of most of the variants including omicron.(23) We have recently shown that T-cell immune responses are largely preserved against the delta variant and offer protection despite decreased neutralising ability of the vaccinated plasma.(2) The final impact on the hospitalisation and mortality rates of the omicron variant-led infections will be dependent on viral factors such as the inherent pathogenicity of the variant and immune evasiveness, host factors such as innate and cellular immune responses, and epidemiological factors such as the proportion of individuals who might have a hybrid immunity from vaccine and past infection. Except for immune evasiveness, the other factors should favour a reduced incidence of severe COVID-19 from the omicron led surge as compared to the previous surges.(24,25)

From a policy perspective, a significant correlation between serum IgG titres and neutralisation GMT against the omicron in the face of immune escape by this variant would lend support for an additional dose of vaccine to augment antibody response. This is being implemented in a few countries including India and has the potential to offer better protection in vulnerable people after 6 months of vaccination or natural infection.

## Data Availability

All data produced in the present study are available upon reasonable request to the authors after approval of national authorities

## Figure Legend

**Figure 1:** Virus neutralization assay: Plasma samples from indicated groups were used for focus reduction neutralization assays with indicated virus strains (N=80). The dotted line represents FRNT50 of 1:20, which was considered as lower limit of quantification and protection.

**Figure 2.** Correlation between FRNT50 titre against ancestral virus and omicron variant, and anti-RBD IgG ELISA levels. The shaded region denotes the 95% confidence intervals.

## Acknowledgment

The authors gratefully acknowledge the DBT Consortium for COVID-19 Research for enabling this study. We sincerely thank participants of the study for providing the blood samples. We thank Dr. Leo Poon from University of Hong Kong for providing the Omicron virus.

## Role of Funding Source

Funding agencies did not have any role in the conception or conduct of the study.

None of the authors were paid by any pharmaceutical company or any other agency to write this article.

All authors had full access to the full data in the study and accept responsibility to submit for publication.

## Author contribution

PKG conceptualized and designed the overall study. SB, RT and NW designed the cohort study; DM, RT, SS, MG, PK, AP, NK enrolled the participants, collected the clinical information and biospecimens. GRM, JS, AA, HS and KP developed and conducted the FRNT experiments; SC, FM, SJD, GB developed and performed the ELISA for antibody titres; GM, RT, DM and AP managed and analysed the clinical and laboratory data; PKG, SB, GRM, GB, RT reviewed and interpreted the data, and wrote the manuscript; all authors reviewed and approved the final manuscript.

## References

1. COVID-19 Map [Internet]. Johns Hopkins Coronavirus Resource Center. [cited 2022 Jan 3]. Available from: https://coronavirus.jhu.edu/map.html

2. Thiruvengadam R, Awasthi A, Medigeshi G, Bhattacharya S, Mani S, Sivasubbu S, et al. Effectiveness of ChAdOx1 nCoV-19 vaccine against SARS-CoV-2 infection during the delta (B.1.617.2) variant surge in India: a test-negative, case-control study and a mechanistic study of post-vaccination immune responses. Lancet Infect Dis. 2021 Nov 25;S1473-3099(21)00680-0.

3. Cromer D, Steain M, Reynaldi A, Schlub TE, Wheatley AK, Juno JA, et al. Neutralising antibody titres as predictors of protection against SARS-CoV-2 variants and the impact of boosting: a meta-analysis. The Lancet Microbe. 2022 Jan 1;3(1):e52–61.

4. Public Health England. SARS-CoV-2 variants of concern and variants under investigation - Technical briefing 29. :45. 26 November 2021 https://assets.publishing.service.gov.uk/government/uploads/system/uploads/attachment_data/file/1036501/Technical_Briefing_29_published_26_November_2021.pdf

5. Cele S, Jackson L, Khoury DS, Khan K, Moyo-Gwete T, Tegally H, et al. SARS-CoV-2 Omicron has extensive but incomplete escape of Pfizer BNT162b2 elicited neutralization and requires ACE2 for infection. medRxiv. 2021 Dec 17;2021.12.08.21267417.

6. Dejnirattisai W, Shaw RH, Supasa P, Liu C, Stuart AS, Pollard AJ, et al. Reduced neutralisation of SARS-CoV-2 omicron B.1.1.529 variant by post-immunisation serum. The Lancet [Internet]. 2021 Dec 20 [cited 2021 Dec 27];0(0). Available from: https://www.thelancet.com/journals/lancet/article/PIIS0140-6736(21)02844-0/fulltext

7. Jahan N, Brahma A, Kumar MS, Bagepally BS, Ponnaiah M, Bhatnagar T, et al. Seroprevalence of IgG antibodies against SARS-CoV-2 in India, March 2020-August 2021: a systematic review and meta-analysis. International Journal of Infectious Diseases [Internet]. 2021 Dec 28 [cited 2022 Jan 4]; Available from: https://www.sciencedirect.com/science/article/pii/S1201971221012510

8. Oeser C, Whitaker H, Linley E, Borrow R, Tonge S, Brown CS, et al. Large increases in SARS-CoV-2 seropositivity in children in England: Effects of the delta wave and vaccination. J Infect [Internet]. 2021 Nov 30 [cited 2022 Jan 5]; Available from: https://www.ncbi.nlm.nih.gov/pmc/articles/PMC8631044/

9. Dhar MS, Marwal R, Vs R, Ponnusamy K, Jolly B, Bhoyar RC, et al. Genomic characterization and epidemiology of an emerging SARS-CoV-2 variant in Delhi, India. Science. 0(0):eabj9932.

10. Mehdi F, Chattopadhyay S, Thiruvengadam R, Yadav S, Kumar M, Sinha SK, et al. Development of a Fast SARS-CoV-2 IgG ELISA, Based on Receptor-Binding Domain, and Its Comparative Evaluation Using Temporally Segregated Samples From RT-PCR Positive Individuals. Front Microbiol. 2020;11:618097.

11. Bewley KR, Coombes NS, Gagnon L, McInroy L, Baker N, Shaik I, et al. Quantification of SARS-CoV-2 neutralizing antibody by wild-type plaque reduction neutralization, microneutralization and pseudotyped virus neutralization assays. Nat Protoc. 2021 Jun;16(6):3114–40.

12. Gu H, Krishnan P, Ng DYM, Chang LDJ, Liu GYZ, Cheng SSM, et al. Early Release - Probable Transmission of SARS-CoV-2 Omicron Variant in Quarantine Hotel, Hong Kong, China, November 2021 - Volume 28, Number 2—February 2022 - Emerging Infectious Diseases journal - CDC. [cited 2022 Jan 4]; Available from: https://www.nc.cdc.gov/eid/article/28/2/21-2422_article

13. Khoury DS, Cromer D, Reynaldi A, Schlub TE, Wheatley AK, Juno JA, et al. Neutralizing antibody levels are highly predictive of immune protection from symptomatic SARS-CoV-2 infection. Nat Med. 2021 Jul;27(7):1205–11.

14. Lu L, Mok BW-Y, Chen L-L, Chan JM-C, Tsang OT-Y, Lam BH-S, et al. Neutralization of SARS-CoV-2 Omicron variant by sera from BNT162b2 or Coronavac vaccine recipients. Clin Infect Dis. 2021 Dec 16;ciab1041.

15. Schmidt F, Muecksch F, Weisblum Y, Da Silva J, Bednarski E, Cho A, et al. Plasma Neutralization of the SARS-CoV-2 Omicron Variant. New England Journal of Medicine. 2021 Dec 30;0(0):null.

16. Rössler A, Riepler L, Bante D, Laer D von, Kimpel J. SARS-CoV-2 B.1.1.529 variant (Omicron) evades neutralization by sera from vaccinated and convalescent individuals [Internet]. 2021 Dec [cited 2022 Jan 4] p. 2021.12.08.21267491. Available from: https://www.medrxiv.org/content/10.1101/2021.12.08.21267491v1

17. Wilhelm A, Widera M, Grikscheit K, Toptan T, Schenk B, Pallas C, et al. Reduced Neutralization of SARS-CoV-2 Omicron Variant by Vaccine Sera and monoclonal antibodies [Internet]. 2021 Dec [cited 2022 Jan 4] p. 2021.12.07.21267432. Available from: https://www.medrxiv.org/content/10.1101/2021.12.07.21267432v1

18. Ai J, Zhang H, Zhang Y, Lin K, Zhang Y, Wu J, et al. Omicron variant showed lower neutralizing sensitivity than other SARS-CoV-2 variants to immune sera elicited by vaccines after boost. Emerg Microbes Infect. 2021 Dec 22;1–24.

19. Khan K, Karim F, Cele S, San JE, Lustig G, Tegally H, et al. Omicron infection enhances neutralizing immunity against the Delta variant [Internet]. 2021 Dec [cited 2022 Jan 2] p. 2021.12.27.21268439. Available from: https://www.medrxiv.org/content/10.1101/2021.12.27.21268439v1

20. Cameroni E, Bowen JE, Rosen LE, Saliba C, Zepeda SK, Culap K, et al. Broadly neutralizing antibodies overcome SARS-CoV-2 Omicron antigenic shift. Nature [Internet]. 2021 Dec 23 [cited 2022 Jan 5]; Available from: https://www.nature.com/articles/d41586-021-03825-4

21. Krutikov M, Palmer T, Tut G, Fuller C, Azmi B, Giddings R, et al. Prevalence and duration of detectable SARS-CoV-2 nucleocapsid antibodies in staff and residents of long-term care facilities over the first year of the pandemic (VIVALDI study): prospective cohort study in England. The Lancet Healthy Longevity [Internet]. 2021 Dec 16 [cited 2022 Jan 4]; Available from: https://www.sciencedirect.com/science/article/pii/S2666756821002828

22. Sette A, Crotty S. Adaptive immunity to SARS-CoV-2 and COVID-19. Cell. 2021 Feb;184(4):861– 80.

23. Keeton R, Tincho MB, Ngomti A, Baguma R, Benede N, Suzuki A, et al. SARS-CoV-2 spike T cell responses induced upon vaccination or infection remain robust against Omicron [Internet]. 2021 Dec [cited 2022 Jan 2] p. 2021.12.26.21268380. Available from: https://www.medrxiv.org/content/10.1101/2021.12.26.21268380v1

24. Zhao H, Lu L, Peng Z, Chen L-L, Meng X, Zhang C, et al. SARS-CoV-2 Omicron variant shows less efficient replication and fusion activity when compared with delta variant in TMPRSS2-expressed cells. Emerg Microbes Infect. 2021 Dec 24;1–18.

25. Public health England. SARS-CoV-2 variants of concern and variants under investigation - Technical briefing 33. 2021 Dec 23;42. https://assets.publishing.service.gov.uk/government/uploads/system/uploads/attachment_data/file/1043807/technical-briefing-33.pdf accessed on January 2, 2022.

