## Supplementary material for "Sub-optimal Neutralisation of Omicron (B.1.1.529) Variant by Antibodies induced by Vaccine alone or SARS-CoV-2 Infection plus Vaccine (Hybrid Immunity) post 6-months": Table S1; Table S2

**Supplementary Materials**

*Quantitative anti-RBD IgG ELISA*

Anti-RBD IgG concentrations in the serum samples were determined by enzyme-linked immunosorbent assay (ELISA) as described earlier with minor modifications required for quantitative output. (1) Internal positive control (pooled human serum from SARS-CoV-2 recovered individuals) was calibrated against the first WHO international standard for anti-SARS-CoV-2 immunoglobulin (code 20/136) as per guidelines (WHO manual for the establishment of national and other secondary standards for antibodies against infectious agents focusing on SARS-CoV-2; Draft Version 12/10/2021) leading to a calculated concentration of 718.3 binding antibody units/ml (BAU/ml) for the positive control. 96-well maxisorp polystyrene plates were coated with RBD antigen, blocked, and dry stabilized as described earlier.(1) Test sera and positive and negative controls were three-fold diluted starting from 1:50 to 1:12150, and 100µl was added to the assay wells. After the incubation, wells were washed, and the bound antibodies were detected using HRP-labelled anti-human IgG (γ-chain specific). Anti-RBD IgG concentrations in the test samples were calculated for each sample dilution by interpolation of OD values on the 4-parameter logistic (4-PL) standard curve from internal positive control using GraphPad Prism 9.3.1 software. Anti-RBD IgG concentrations above the assay cut-off and corresponding to the linear part of the curve were considered, and values in BAU/ml were assigned to each test sample. The lower limit of quantitation for the assay was 24 BAU/ml. Additional dilutions beyond 1:12150 were done for samples where OD values were beyond the calibration curve's linear part.

*Qualitative anti-nucleocapsid IgG ELISA*

We tested for IgG antibodies against SARS-CoV-2 nucleocapsid antigen by ELISA as a measure of past infection with SARS-CoV-2. The assay procedure was similar to the process described for the RBD IgG ELISA(1) with the following modifications: 1) instead of RBD, *E.coli* expressed SARS-CoV-2 nucleocapsid antigen was coated on polystyrene wells (50ng/50µl); 2) sample dilution of 1:200 was used. Samples with a signal/cut-off ratio above 1 were considered positive for anti-nucleocapsid antibodies.

**Supplementary Results**

**Table S1. Anti-nucleocapsid IgG in different groups**

| **Group** | **n** | **Anti-N Positive** |
| --- | --- | --- |
| BBV152 | 20 | 8 |
| BBV152+Infection | 20 | 18 |
| ChAdOx1 nCoV-19 | 20 | 1 |
| ChAdOx1 nCoV-19 +Infection | 20 | 11 |

**Table S2: Anti-RBD IgG titres (BAU/ml) in different groups**

| **Group** | **n** | **Anti-RBD IgG**  **median (IQR)** | **Minimum** | **Maximum** |
| --- | --- | --- | --- | --- |
| All* | 79 | 243.5 (111.3, 544.4) | 27.9 | 7201 |
| BBV152 and BBV152+Infection* | 39 | 226.7 (83.3, 414.3) | 27.9 | 2378 |
| ChAdOx1 nCoV-19  and  ChAdOx1 nCoV-19 +Infection | 40 | 284.9 (134.8, 636.3) | 27.9 | 7201 |
| BBV152 | 20 | 213.3 (57.38, 378.1) | 27.9 | 594.1 |
| BBV152+Infection* | 19 | 301.5 (127.1, 665.0) | 43.2 | 2378 |
| ChAdOx1 nCoV-19 | 20 | 142.4 (99.0, 283.1) | 27.9 | 1160 |
| ChAdOx1 nCoV-19 +Infection | 20 | 541.4 (290.3, 966.3) | 35 | 7201 |
| Vaccine alone | 40 | 150.6 (81.9, 298.5) | 27.9 | 1160 |
| Vaccine + infection* | 39 | 403.9 (164.6, 794.8) | 35 | 7201 |
| Anti-nucleocapsid IgG positive* | 37 | 380.7 (178.0, 729.9) | 35 | 7201 |
| Anti-nucleocapsid IgG Negative | 42 | 161.6 (82.8, 338.4) | 27.9 | 6808 |
| BBV152 and BBV152+Infection Anti- nucleocapsid IgG Positive* | 25 | 301.5 (123.5, 640.1) | 43.2 | 2378 |
| BBV152 and BBV152+Infection Anti- nucleocapsid IgG Negative | 14 | 105.2 (45.1, 281.0) | 27.9 | 414.3 |
| ChAdOx1 nCoV-19  and  ChAdOx1 nCoV-19 +Infection  Anti- nucleocapsid IgG Positive | 12 | 624.8 (231.4, 1146.0) | 35 | 7201 |
| ChAdOx1 nCoV-19  and  ChAdOx1 nCoV-19 +Infection  Anti- nucleocapsid IgG Negative | 28 | 212.1 (112.9, 344.4) | 27.9 | 6808 |

* One of the samples had a very high level of anti-RBD IgG titre (85443.9 BAU/ml) and was considered an outlier and not included in this analysis.

References:

1.	Mehdi F, Chattopadhyay S, Thiruvengadam R, Yadav S, Kumar M, Sinha SK, et al. Development of a Fast SARS-CoV-2 IgG ELISA, Based on Receptor-Binding Domain, and Its Comparative Evaluation Using Temporally Segregated Samples From RT-PCR Positive Individuals. Front Microbiol. 2020;11:618097.
